# CLINICO ETIOLOGICAL PROFILE OF URINARY TRACT INFECTION AND UROLITHIASIS

**DOI:** 10.1101/2025.10.26.25338625

**Authors:** Mohammed Yousuff, R Rajashekar, N Monika, Jaish George

**Affiliations:** DEPARTMENT OF NEPHROLOGY, RAMAIAH MEDICAL COLLEGE AND HOSPITAL

## Abstract

**BACKGROUND:** The prevalence of urolithiasis in the general population has increased worldwide in the past decades. The aim of this study is to investigate the bacteriological profile of urine samples from patients with kidney stones. By analyzing the bacteria present in the urine, we can gain insights into potential associations between specific types of stones and the microbial flora. This information can be crucial for tailoring treatment strategies, including antibiotic therapy and preventive measures.

**METHODOLOGY:** It is a retrospective cross-sectional study comprised of all patients who were diagnosed with urolithiasis at Ramaiah Memorial Hospital, Bengaluru, Karnataka. In this study we assessed a total of 235 patients who were admitted to our hospital due to kidney stones. The socio-demographic and epidemiological profile. Type of calculus, urinalysis and treatment method was considered.

**RESULTS:** Out of 235 patients, 189 (80.4%) were males, and 46 (19.6%) were females. 80 patients (34% of the total patients) were found to have clinical or laboratory evidence of urinary tract infection (UTI). Most common organism isolated was Escherichia coli in 19 patients (48.7%) followed by Klebsiella in 15 patients (38.46%). In biochemical analysis most common type of stone found was calcium oxalate monohydrate (97.43%) and calcium oxalate dihydrate (66.66%).

**CONCLUSION:** Positive urine culture in stone formers should not be assumed to indicate purely infection-related stones; metabolic evaluation remains essential. In patients with recurrent CaOx stones and persistent bacteriuria, targeted antibiotic therapy and biofilm-disrupting strategies could be considered to reduce recurrence.

## INTRODUCTION

Urolithiasis is a term used to describe calculi or stones that form in the urinary system, usually in the kidneys or ureters, but may also affect the bladder or urethra(1). These are formed from the accumulation of substances like calcium, oxalate, cystine or uric acid which are present in urine(2).

The prevalence of urolithiasis in the general population has increased worldwide in the past decades, the recurrence rate of urinary stones is high and is estimated to be approximately 50% at 10 years(3). One of the most typical conditions for which a patient comes to our hospital’s outpatient department is urolithiasis. About 7% to 13% of North Americans, 5% to 9% of Europeans, and 1% to 5% of Asians are affected(4).

The formation of stones is a complex biological process. Urinary solute supersaturation provides the necessary nidus for formation of stones. Super saturation is a process through which the concentration of chemicals in urine, such as calcium and oxalate, exceeds the limits of their solubility. Majority of kidney stones worldwide are made of calcium oxalate (CaOx) and calcium phosphate (CaPhos). Other common stone types include those made of struvite, cysteine, and uric acid, which make up about 9%, 10%, and 1% of all stones, respectively (5).

Bacteria can play a significant role in the formation of urinary tract stones. They can contribute to stone formation by promoting the precipitation of minerals, by providing a surface for mineral deposition, and by causing inflammation and tissue damage.

The aim of this study is to investigate the bacteriological profile of urine samples from patients with kidney stones. By analyzing the bacteria present in the urine, we can gain insights into potential associations between specific types of stones and the microbial flora in the urinary tract. This information can be crucial for tailoring treatment strategies, including antibiotic therapy and preventive measures, to better address the underlying causes of urolithiasis and reduce the risk of recurrence.

Overall, the study aims to contribute to a better understanding of the role of bacteriology in urolithiasis and to inform more effective management approaches for patients with kidney stones.

## METHODOLOGY

It is a retrospective cross-sectional study comprised of all patients who were diagnosed with urolithiasis at Ramaiah Memorial Hospital, Bengaluru, Karnataka. In this study we assessed a total of 235 patients who were admitted to our hospital due to kidney stones. The socio-demographic and epidemiological profile which contained the name, age, sex, caste along with the diagnosis was entered in a file. Type of calculus, urinalysis and treatment method was considered. Data was analysed using **SPSS (Statistical Package For Social Sciences)** version 21. (IBM SPASS statistics [IBM corporation: NY, USA]). Data was entered in the excel spread sheet. Descriptive statistics of the explanatory and outcome variables were calculated by mean, standard deviation for quantitative variables, frequency and proportions for qualitative variables. Inferential statistics like Chi-square test was applied for qualitative variables to find the association. The level of significance is set at 5%.

## RESULTS

### DEMOGRAPHICS

#### SEXWISE DISTRIBUTION

Out of 235 patients, 189 (80.4%) were males, and 46 (19.6%) were females.

**TABLE 1:** DISTRIBUTION OF THE SUBJECTS BASED ON GENDER.

| Gender | Frequency | Percent |
| --- | --- | --- |
| Females | 46 | 19.6 |
| Males | 189 | 80.4 |
| Total | 235 | 100.0 |

#### AGEWISE DISTRIBUTION

The mean age of the patients was 41.54 years, with a standard deviation of ±16.75 years. Majority of the patients were in the age group of 31-40years (23.4%) followed by 51-60 years (19.1%) age group. It was least common among patients aged >70 years (3.8%).

**TABLE 2:** DISTRIBUTION OF THE SUBJECTS BASED ON AGE GROUPS.

| Age Groups | Frequency | Percent |
| --- | --- | --- |
| < 10 yrs | 11 | 4.7 |
| 11 to 20 yrs | 10 | 4.3 |
| 21 to 30 yrs | 40 | 17.0 |
| 31 to 40 yrs | 55 | 23.4 |
| 41 to 50 yrs | 43 | 18.3 |
| 51 to 60 yrs | 45 | 19.1 |
| 61 to 70 yrs | 22 | 9.4 |
| > 70 yrs | 9 | 3.8 |
| Total | 235 | 100.0 |

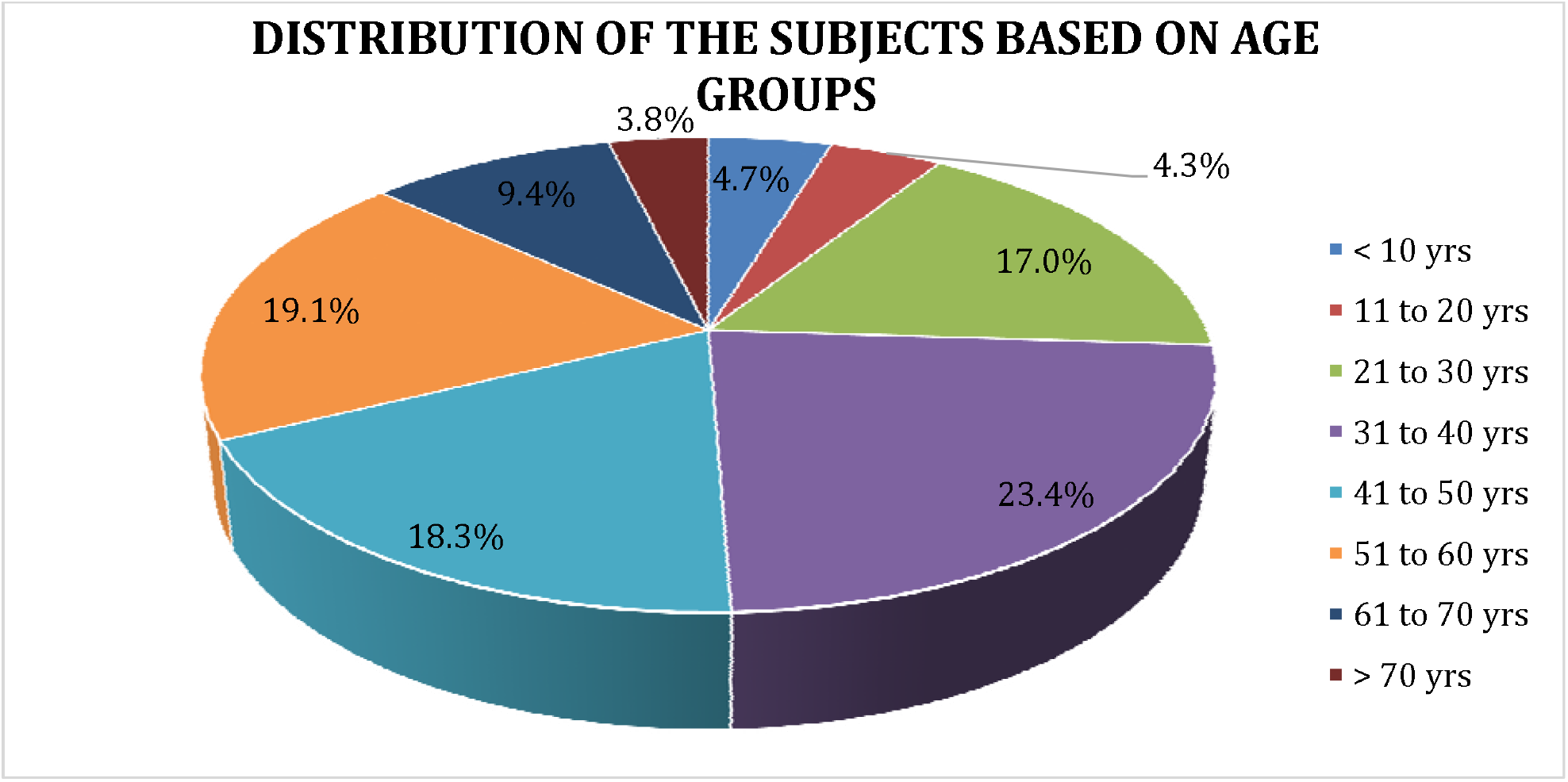

#### COMORBIDITIES

Among the patients assessed, 18 had hypertension and a significant number of patients, 110 in total had Type 2 Diabetes Mellitus.

#### LABORATORY PARAMETERS

Mean values of creatinine, calcium, phosphorous, uric acid were 1.17mg/dl, 8.86mg/dl, 4.05mg/dl, 5.52mg/dl respectively. Mean total leucocyte count was 22,688.

**TABLE 3:**
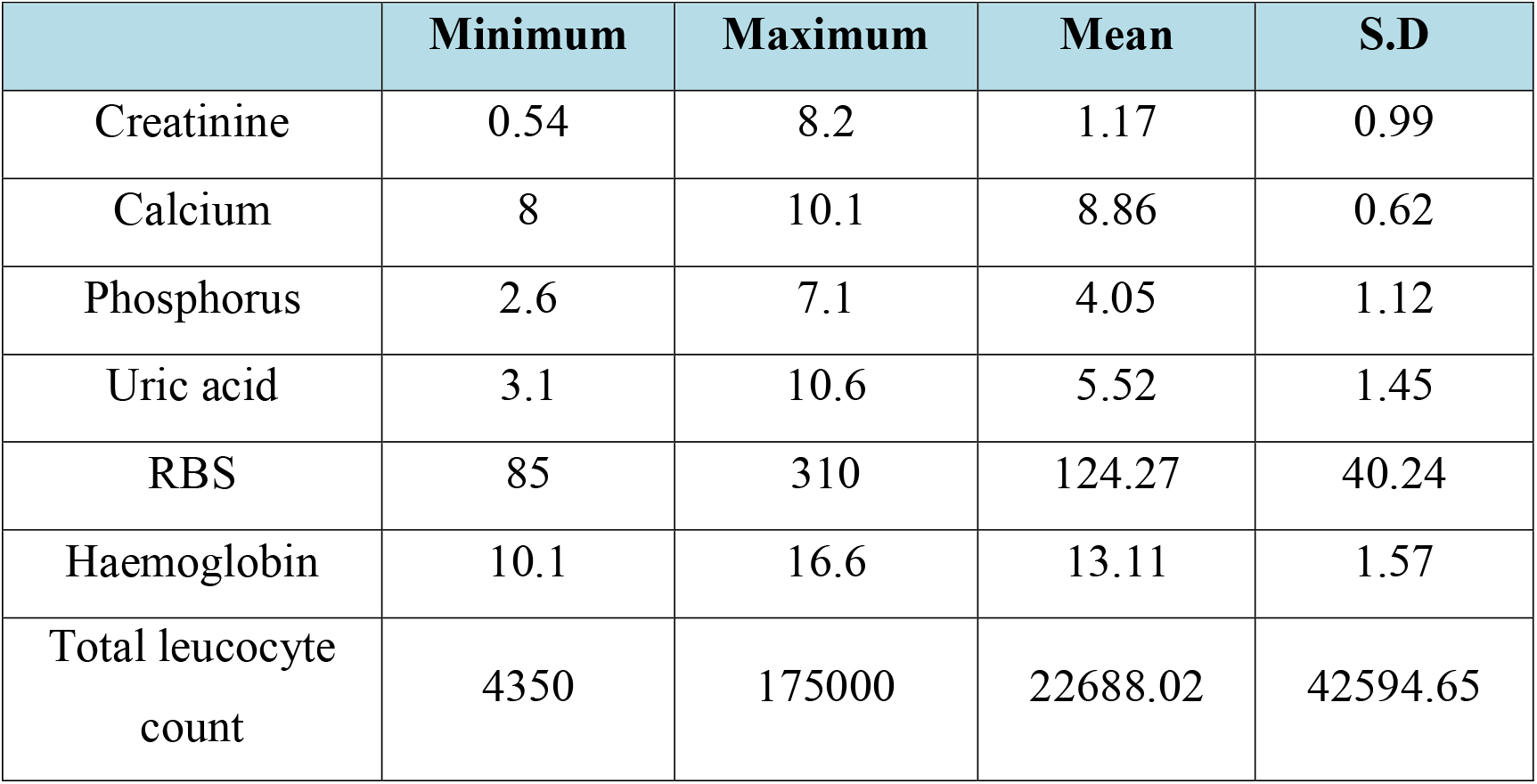
LABORATORY PARAMETERS.

#### URINALYSIS

119(50.6%) of the patients had urine albumin 1+ by dipstick method. (38)16.2% of the patients had urine albumin 2+ and 2(0.9%) patients had 3+ albuminuria. 76(32.3%) patients had no proteinuria.

**TABLE 4:** DISTRIBUTION OF THE SUBJECTS BASED ON URINE ALBUMIN.

| Urine Albumin | Frequency | Percent |
| --- | --- | --- |
| 1+ | 119 | 50.6 |
| 2+ | 38 | 16.2 |
| 3 | 2 | .9 |
| Nil | 76 | 32.3 |
| Total | 235 | 100.0 |

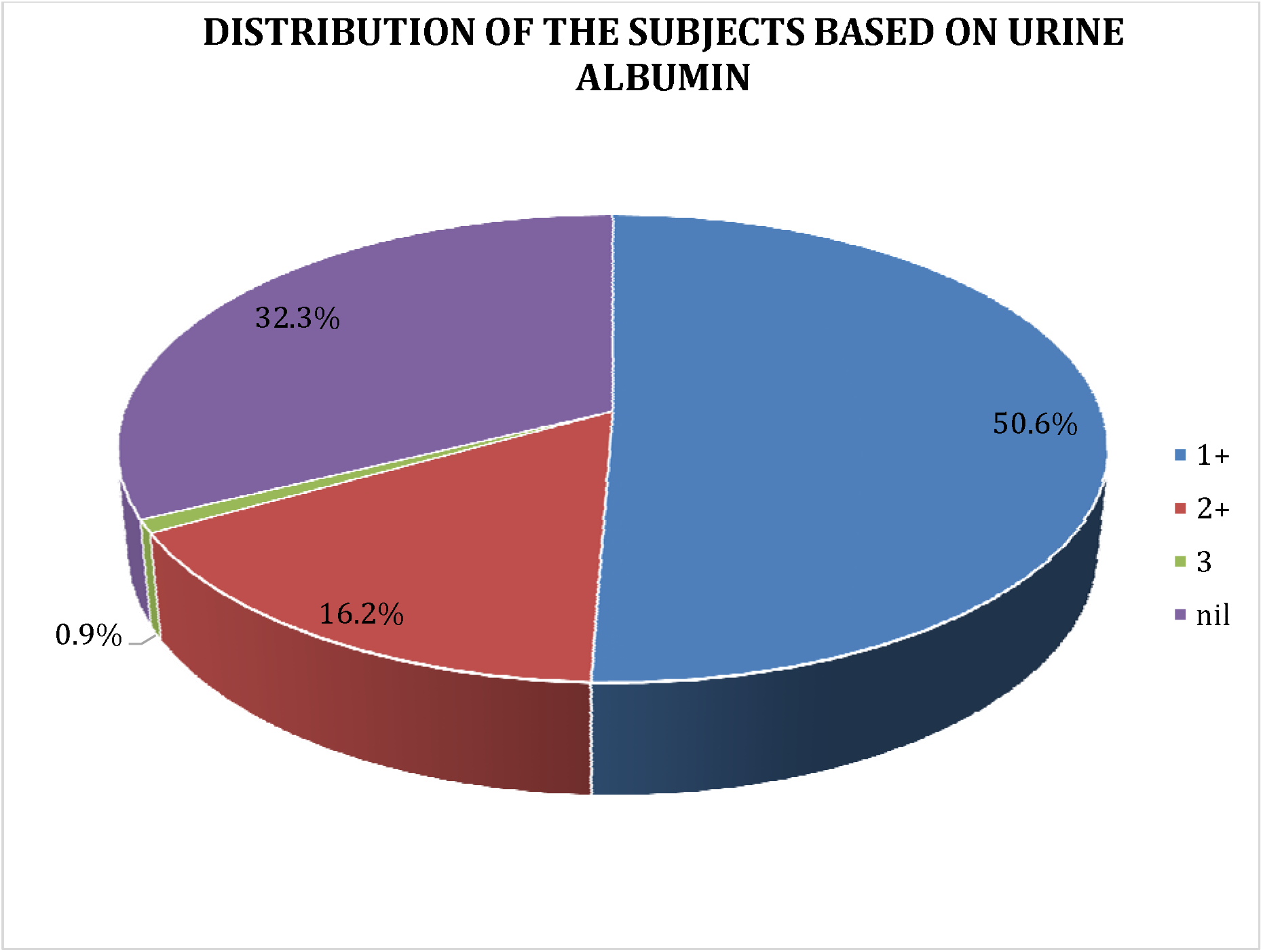

80 patients (34% of the total patients) were found to have clinical or laboratory evidence of urinary tract infection (UTI), characterized by the presence of pus cells in urine (>5 per high-power field). Among these 80 patients with clinical/laboratory UTI, 39(48.7%) showed bacterial growth in urine culture. Most common organism isolated was Escherichia coli in 19 patients (48.7%) followed by Klebsiella in 15 patients (38.46%). Among E.Coli positive urine cultures, 16 (84.2%) samples were sensitive to Piperacillin tazobactum. Among Klebsiella positive cultures, majority were sensitive to Meropenem (66.6%). Panresistance pattern was seen in about 5(33.3%) patients with Klebsiella positive cultures. Additionally, out of the total patient population, 110 were diabetic. Among the diabetic patients, 30 had positive urine cultures indicating the presence of UTI. Interestingly, one patient who did not have pus cells in the urine sample showed positive bacterial growth in the urine culture.

**TABLE 5:**
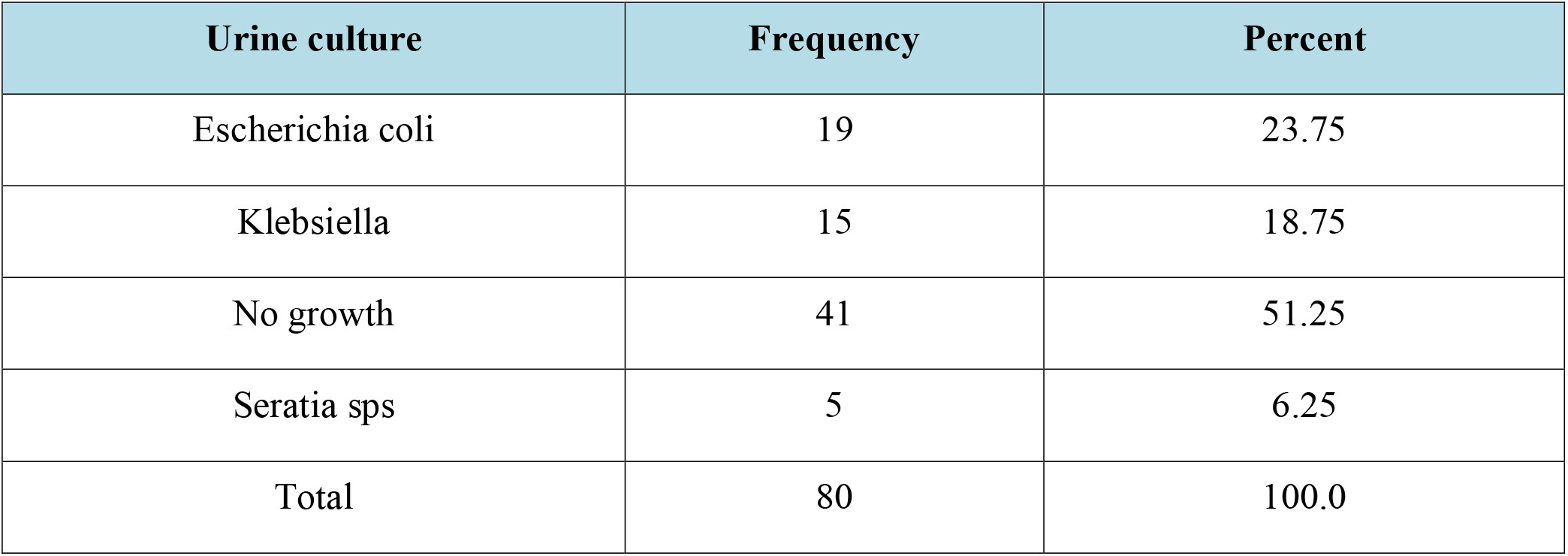
DISTRIBUTION OF THE SUBJECTS BASED ON URINE CULTURE.

| Urine culture | Frequency | Percent |
| --- | --- | --- |
| Escherichia coli | 19 | 23.75 |
| Klebsiella | 15 | 18.75 |
| No growth | 41 | 51.25 |
| Serratia sps | 5 | 6.25 |
| Total | 80 | 100.0 |

**TABLE 6:**
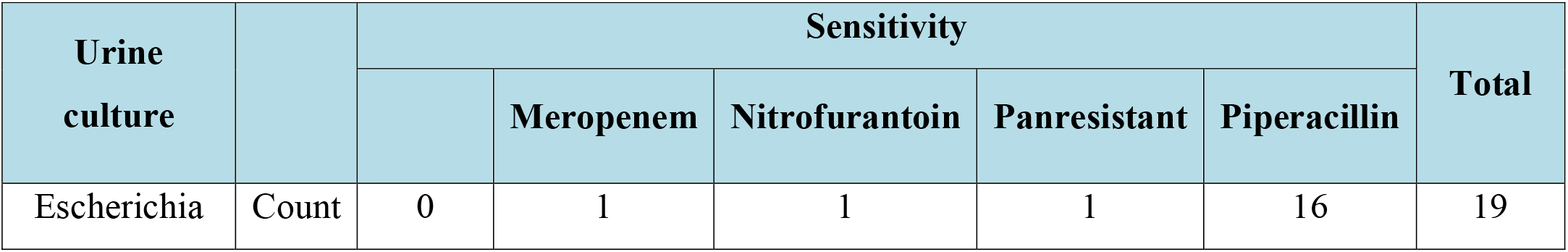
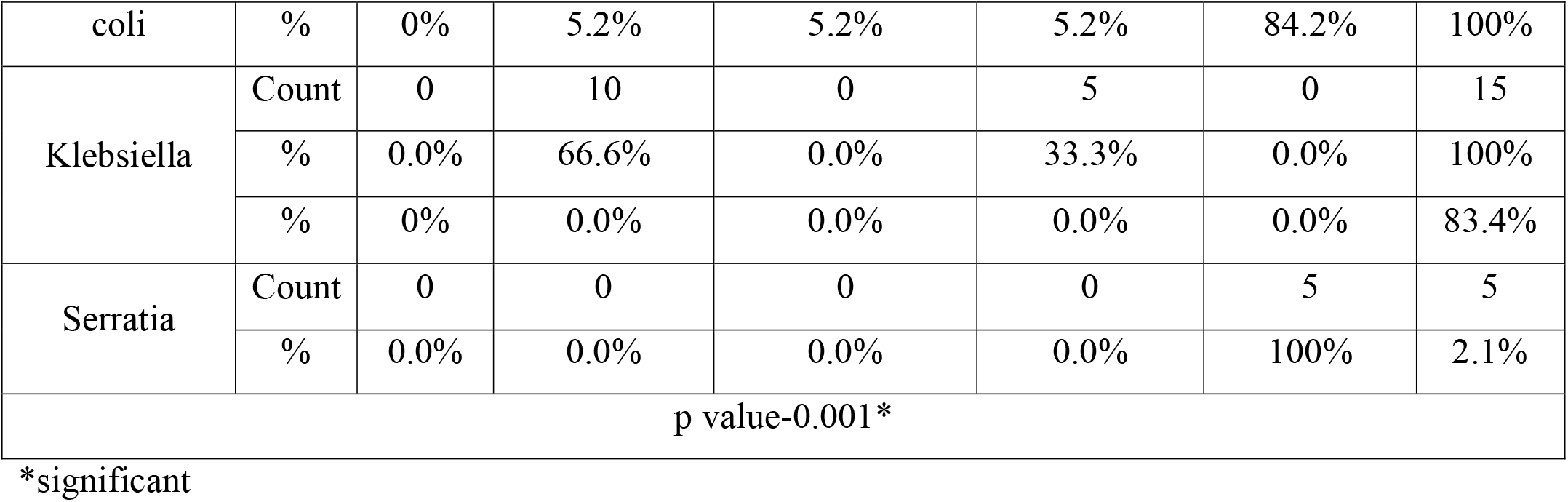
CROSS-TABULATION OF URINE CULTURE AND SENSITIVITY.

| Urine culture |  | Sensitivity |  |  |  |  | Total |
| --- | --- | --- | --- | --- | --- | --- | --- |
|  |  |  | Meropenem | Nitrofurantoin | Panresistant | Piperacillin |  |
| Escherichia | Count | 0 | 1 | 1 | 1 | 16 | 19 |
| coli | % | 0% | 5.2% | 5.2% | 5.2% | 84.2% | 100% |
| Klebsiella | Count | 0 | 10 | 0 | 5 | 0 | 15 |
|  | % | 0.0% | 66.6% | 0.0% | 33.3% | 0.0% | 100% |
|  | % | 0% | 0.0% | 0.0% | 0.0% | 0.0% | 83.4% |
| Serratia | Count | 0 | 0 | 0 | 0 | 5 | 5 |
|  | % | 0.0% | 0.0% | 0.0% | 0.0% | 100% | 2.1% |
| p value-0.001* |  |  |  |  |  |  |  |

\*significant

#### SITE OF STONE FORMATION

Left ureter was the most common site of calculi formation (28.9%) followed by right renal calculi (23.4%).

**TABLE 7:** DISTRIBUTION OF THE SUBJECTS BASED ON SITE OF STONE.

| Site of Stone | Frequency | Percent |
| --- | --- | --- |
| B/L renal calculi | 32 | 13.6 |
| Left renal calculi | 33 | 14.0 |
| Left Ureteric calculi | 68 | 28.9 |
| Left ureteric calculi with B/L renal calculi | 2 | .9 |
| Left Ureteric calculus with left Renal calculus | 1 | .4 |
| Non - obstructive right renal calculus | 2 | .9 |
| Right renal calculi | 55 | 23.4 |
| Right renal calculi/right ureteric calculi | 4 | 1.7 |
| Right ureteric calculi | 38 | 16.1 |
| Total | 235 | 100.0 |

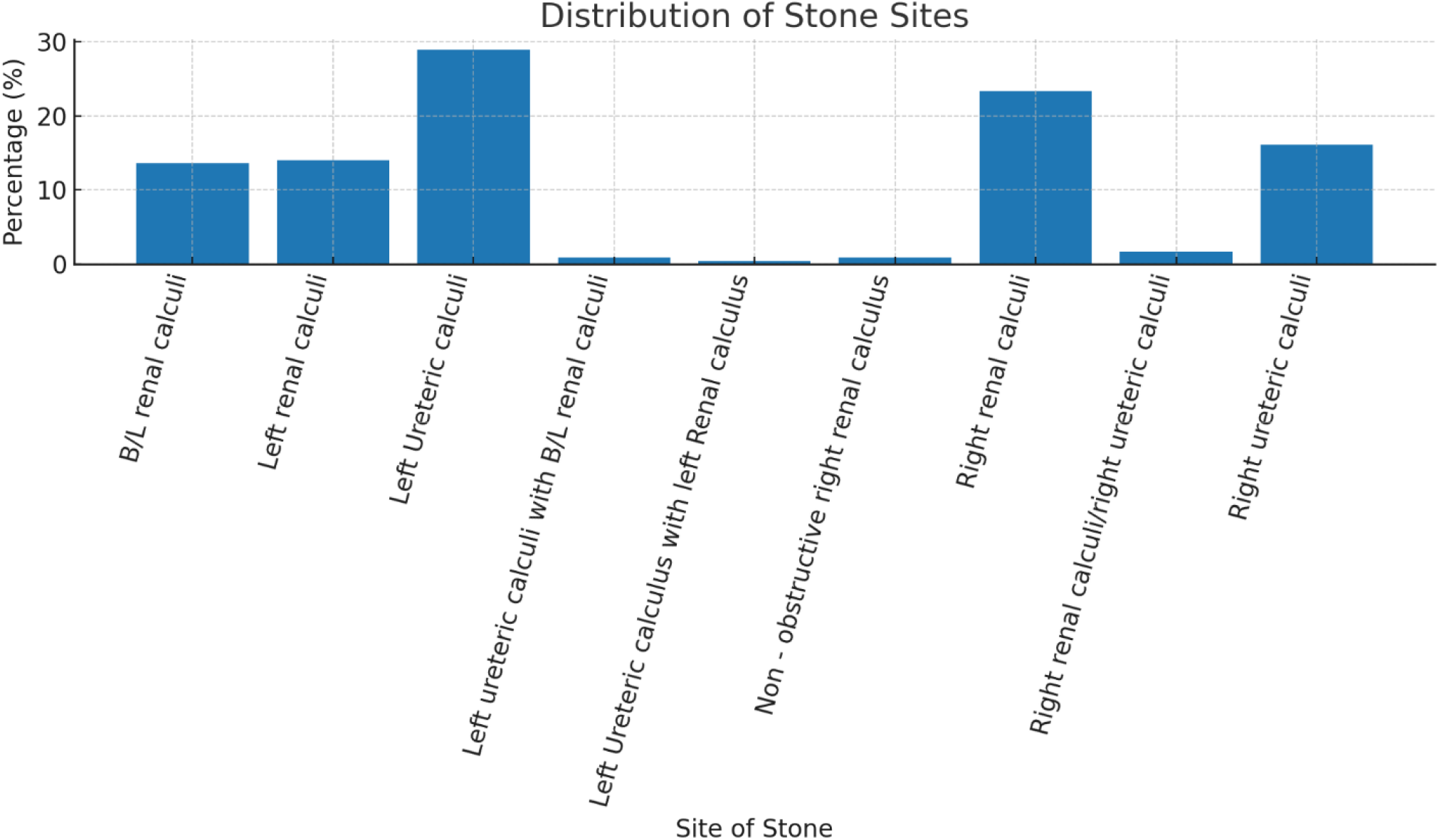

In biochemical analysis most common type of stone found was calcium oxalate monohydrate (97.43%) and calcium oxalate dihydrate (66.66%), other constituents found in stones are ammonium magnesium phosphate, carbonate apatite, anhydrous uric acid, Amorphous carbonated Ca-phosphate, Ammonium urate, uric acid dihydrate, sodium urate, Ammonium magnesium phosphate hexahydrate.

Most predominantly 12 culture positive patients had more than 2 components in the stone analysis, 68 patients who have no culture growth had Calcium oxalate monohydrate and calcium oxalate dihydrate in the stone analysis, 17 patients with positive culture growth had CaoX in the stones.

### MANAGEMENT

DJ stenting was the most commonly performed procedure (38.7%) followed by URSL (28.1%).

**TABLE 12:** DISTRIBUTION OF THE SUBJECTS BASED ON PROCEDURES.

| Site of Stone | Frequency | Percent |
| --- | --- | --- |
| DJ stent | 91 | 38.7 |
| NIL | 12 | 5.1 |
| PCNL | 31 | 13.2 |
| PCNL+DJ stent | 2 | .9 |
| RIRS | 30 | 12.8 |
| RIRS +DJ stent | 1 | .4 |
| URSL | 66 | 28.1 |
| URSL+DJ stent | 2 | .9 |
| Total | 235 | 100.0 |

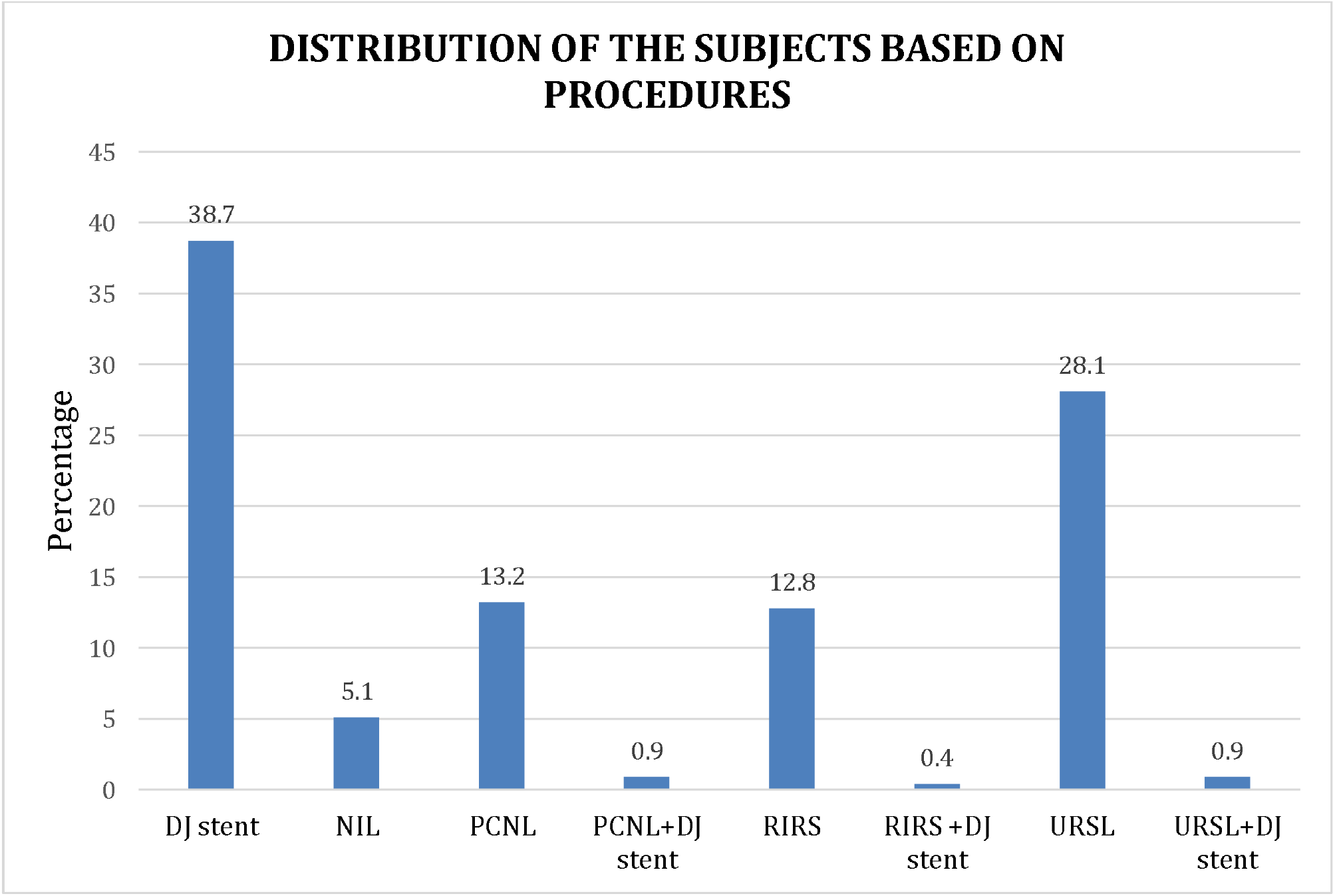

## DISCUSSION

This observational study was conducted with 235 patients who came to our hospital with kidney stones. The patients were assessed with preoperative urine culture and postoperative kidney stones were analyzed. In this study majority of patients were male (189) compared to female (46) which was similar to earlier studies by different authors that suggested that men were predominant stone formers. Men may experience symptomatic stone illness up to three times more frequently than women, according to US health care data(6). At a rate of 8:1, females get UTIs more frequently than males do. The male predominance can be explained by the role of androgens in enhancing the oxalate excretion and deposition of calcium in the urinary system whereas in female, estrogen decrease oxalate excretion(7). Therefore, there is more burden of stones among males than in females. Patients with urolithiasis frequently experience UTI as a consequence. A major cause of urinary tract blockage that increases the risk of UTI is urolithiasis(8).

Overall, the incidence of urinary tract stones increased with age, which peaked in the age group of 30–60 years and decreased afterwards(9). The reason why middle-age population are prone to urolithiasis is because they do more laborious work than others, and subsequently result in less fluid intake and higher rate of dehydration. Unhealthy lifestyle and occupational stress also adds to the risk factors(9).

In our study, 34% of patients had clinical or laboratory evidence of urinary tract infection (UTI), and 16.6% of the total cohort had a positive urine culture. The most common organisms isolated were *Escherichia coli* (48.7%), followed by *Klebsiella* spp. (38.4%) and *Serratia* spp. (12.8%). Interestingly, the predominant stone composition in culture-positive patients was calcium oxalate (CaOx), despite the conventional understanding that infection-related stones are usually struvite or carbonate apatite(10,11).

### Correlation between Positive Urine Culture and Stone Type

Among culture-positive patients, **CaOx monohydrate and dihydrate** remained the most common constituents, mirroring the culture-negative group. Struvite stones, classically associated with urease-producing organisms like *Proteus* spp., were not the most frequent type in our culture-positive patients. This is likely because *E. coli*, the predominant organism in our cohort, is not a potent urease producer(12), although certain strains have been reported to possess weak urease activity and other virulence factors facilitating biofilm formation(13). *Klebsiella* spp., isolated in 38.4% of culture-positive patients, is a urease producer and could contribute to the pathogenesis of struvite stones(14); however, in our dataset, mixed-composition stones (e.g., CaOx + struvite) were more frequent than pure struvite stones. *Serratia* spp., though less common, are also urease producers and have been implicated in infection-related stone formation, suggesting that even small proportions of such organisms can influence stone composition(15).

### Possible Pathophysiological Links

1. **Biofilm Formation:** Uropathogens like *E. coli* and *Klebsiella* can form biofilms on urothelium and existing crystals, acting as a nidus for mineral deposition(13,16).
2. **Mixed Stone Composition:** Many culture-positive patients had >2 stone components, suggesting that infection can superimpose on pre-existing metabolic stone disease.
3. **Diabetes as a Risk Factor:** A significant proportion (27.3%) of diabetics in our cohort had positive urine cultures, supporting the known link between impaired immunity, glycosuria, and bacterial colonization(17).

### Comparison with Literature

Previous studies 【3,16,18】 have demonstrated that bacterial DNA can be isolated from calcium oxalate stones, suggesting that even non-urease-producing organisms can be involved in stone pathogenesis via adhesion and aggregation mechanisms. Our findings support this hypothesis, given the high prevalence of *E. coli* in CaOx stones in culture-positive patients.

### Clinical Implications

- Positive urine culture in stone formers should not be assumed to indicate purely infection-related stones; metabolic evaluation remains essential.
- In patients with recurrent CaOx stones and persistent bacteriuria, targeted antibiotic therapy and biofilm-disrupting strategies could be considered to reduce recurrence.
- Mixed-composition stones in culture-positive patients suggest the need for both infection control and metabolic correction.

## Data Availability

All data produced in the present work are contained in the manuscript

